# Prevalence of and risk factors for peripheral artery disease in rural South Africa: A cross-sectional analysis of the HAALSI cohort

**DOI:** 10.1101/2023.07.18.23292814

**Authors:** Erika Teresa Beidelman, Molly Rosenberg, Alisha N. Wade, Nigel Crowther, Corey A. Kalbaugh

## Abstract

**Introduction:** The burden of peripheral artery disease (PAD) is increasing in low- and middle-income countries. Existing literature from sub-Saharan Africa is limited and lacks population-representative estimates. We estimated the burden and risk factor profile of PAD for a rural South African population.

**Methods:** We used data from 1,883 participants of the HAALSI cohort of South African adults aged 40-69 years with available ankle-brachial index (ABI) measurements. We defined clinical PAD as ABI ≤0.90 or >1.40 and borderline PAD as ABI >0.90 & ≤1.00. We compared the distribution of sociodemographic variables, biomarkers, and comorbidities across PAD classifications. To identify associated factors, we calculated unadjusted and age-sex-adjusted prevalence ratios with log-binomial models.

**Results:** Overall, 6.6% (95% CI: 5.6-7.7) of the sample met the diagnostic criteria for clinical PAD while 44.7% (95% CI: 42.4-47.0) met the diagnostic criteria for borderline PAD. Age (PR: 1.9, 95% CI: 1.2-3.1 for ages 50-59 compared to 40-49; PR: 2.5, 95% CI: 1.5-4.0 for ages 60-69 compared to 40-49) and C-reactive protein (PR: 1.08, 95% CI: 1.03-1.12) were associated with increased prevalence of clinical PAD. All other examined factors were not associated with clinical PAD.

**Conclusions:** We found high PAD prevalence for younger age groups compared to previous research and a lack of evidence for the influence of traditional risk factors for this rural, low-income population. Future research should focus on identifying the underlying risk factors for PAD in this setting. South African policymakers and clinicians should consider expanded screening for early PAD detection in rural areas.

## Introduction

People in low- and middle-income countries (LMICs) are living longer, and the incidence of important chronic diseases associated with aging is rapidly increasing ^1,2^. People in LMICs are also more likely to live in rural settings than people in high income countries and this rurality causes additional resource challenges ^3^. Within this context of ongoing demographic and disease burden transitions across LMICs, it is critical to better manage preventable conditions to reduce healthcare costs, reduce the burden on resource-constrained health systems, and promote healthy longevity.

One chronic disease that is important in LMICs is peripheral artery disease (PAD), an atherosclerotic disorder of the lower extremities that contributes to reduced quality of life as well as increased risk of adverse cardiovascular events and premature death ^4–6^. In fact, 73% of the global burden of PAD is concentrated in LMICs with particularly high prevalence estimates in sub-Saharan Africa where traditional risk factors, such as diabetes and hypertension, are known to be poorly controlled ^7,8^. Interestingly, there is also evidence that the risk factor profile for PAD may differ in sub-Saharan African populations compared to populations in high-income countries, making PAD more difficult to predict and, thus, more difficult to identify and manage. For example, in sub-Saharan Africa, PAD prevalence is significantly higher among females compared to males ^9–12^. In contrast, few gender differences have been observed for PAD prevalence in high-income countries ^8^. Despite this preliminary evidence, the existing literature for PAD in sub-Saharan Africa is from clinical samples and disease modeling efforts, representing significant limitations in the literature ^8–14^. Thus, further research in sub-Saharan African countries is needed to better understand the burden and risk profile for PAD.

For PAD, rural South Africa is an understudied location with an important distribution of sociodemographic and comorbid risk factors such as high HIV prevalence, low socioeconomic status, and high burden of multi-morbid chronic diseases ^15–17^. We used data from the “Health and Aging in Africa: A Longitudinal Study of an INDEPTH Community” (HAALSI) cohort, a population-representative cohort of rural, low-income South Africans aged 40 years and older to quantify the burden and risk factor profile of PAD. This analysis contributes to the literature as the first PAD burden study specific to rural South Africans.

## Methods

### Dataset and Source Population

This analysis used data from the HAALSI cohort study of older adults in rural South Africa. The HAALSI cohort was recruited in Agincourt, South Africa and includes three waves of data collection (2014/15, 2017/18, 2019/21). The HAALSI cohort is nested within the Agincourt Health and Demographic Surveillance System (HDSS), which maintains a research platform that fully enumerates the population living across 31 villages (population 118,000+) in rural Mpumalanga Province ^18^. In 2014, potential participants were randomly selected for invitation to join the cohort from a sampling frame of all individuals in the study area greater than or equal to 40 years of age ^16^. The complete HAALSI cohort consisted of 5059 men and women at baseline ^16^. Following enrollment and consent, household respondents participated in baseline in-person interview surveys, physical measurements, and voluntary point-of-care biomarker testing ^16^. A sub-sample of 1937 participants below the age of 70 years participated in supplemental data collection that included additional cognitive, physical, and laboratory measurements ^19^.

The Agincourt research site served by the Agincourt HDSS is a former homeland area where Black Africans were forcibly moved under the apartheid policy ^20^. This area has high rates of unemployment, high fertility rates, high HIV prevalence, and low access to key infrastructure compared to the general population ^16,20^. The HAALSI study has ethical approvals from the Mpumalanga Provincial Research and Ethics Committee, the University of the Witwatersrand Human Research Ethics Committee (ref. M141159), and the Harvard T.H. Chan School of Public Health Office of Human Research Administration (ref. C13–1608–02) ^16^. Ethical approvals were also sought from the Indiana University Institutional Review Board for this analysis where it was deemed not human subjects research because analysis was conducted on deidentified datasets (ref. 18808).

### Study Sample

We used HAALSI baseline data collected between November 2014 and November 2015 as well as HAALSI’s baseline laboratory dataset to perform this cross-sectional analysis. To be included in this study, participants must have participated in the baseline household survey and selected for and participated in the supplemental laboratory testing. Only participants younger than age 70 were eligible for the supplemental laboratory testing. We identified 1883 participants aged 40-69 years for inclusion based on having available ankle-brachial index (ABI) data. Fifty-four individuals were excluded from our analysis because they did not have sufficient data to calculate an ABI.

### Peripheral Artery Disease

PAD was the primary outcome of interest for this study. We identified cases of clinical and borderline PAD among participants with the ankle-brachial index (ABI). ABI is the most common, noninvasive diagnostic measure for PAD and represents the ratio of systolic blood pressure in an individual’s ankle to their brachial artery ^5,6^. During HAALSI’s supplemental laboratory testing, two systolic blood pressure measurements were recorded in millimeters of mercury (mmHg) for each of the following arteries: left dorsalis pedis, right dorsalis pedis, left posterior tibial, right posterior tibial, left brachial, and right brachial. An average of the two measurements was calculated for each artery. Then to calculate the ABI, three steps were sequentially completed for both the left and right leg: (1) We identified the higher systolic blood pressure of the two leg measurements (dorsalis pedis or posterior tibial); (2) Analysts divided the leg measurement by the higher of two brachial systolic blood pressures (right or left); and (3) Analysts recorded the lower ABI value of the two legs (right or left) to two decimal places ^21^.

In line with consensus guidelines from the American Heart Association, clinical PAD was defined as an ABI of less than or equal to 0.90 or greater than 1.40 while borderline PAD was defined as an ABI greater than 0.90 and less than or equal to 1.00 ^4–6^. Participants were considered to have no indications of PAD if their ABI was greater than 1.00 and less than or equal to 1.40. Binary indicator variables were coded for both clinical PAD and borderline PAD based on the above definitions. Cases were coded as ‘1’ for each variable and non-cases were coded as ‘0’.

### Comorbidities & Sociodemographic Characteristics

We examined a set of sociodemographic characteristics, biomarker measures, and comorbidities to characterize the population and examine traditional risk factors that have been established within the existing literature ^4,5,22^. Sociodemographic characteristics used in our analyses included gender (male vs. female), age (in years), education level (none, primary or some secondary, vs. secondary or higher), place of birth (within South Africa vs. outside South Africa), prior occupational smoke exposure (yes vs. no), smoking status (current smoker, ever-smoker, vs. non-smoker), and current alcohol drinking status (yes vs. no). All sociodemographic variables were collected via self-report during the HAALSI baseline in-person survey.

Biomarker measures used in our analyses included high-density lipoprotein (HDL) cholesterol (in millimoles per liter (mmol/L)), low-density lipoprotein (LDL) cholesterol (in mmol/L), total cholesterol (in mmol/L), triglycerides (in mmol/L), blood glucose (in mmol/L), diastolic blood pressure (in mmHg), systolic blood pressure (in mmHg), hemoglobin (in grams per deciliter (g/dL)), and C-reactive protein (CRP) (in milligrams per liter (mg/L)). All biomarker measures were taken during the baseline in-person interview via point-of-care blood testing, dried blood spot testing, and blood pressure testing, using previously described methodologies ^23^.

We assessed the following comorbidities among participants: HIV status, diabetes, dyslipidemia, hypertension, obesity, prior adverse cardiac event, and kidney disease. Binary variables for HIV status, prior adverse cardiac events, and kidney disease were identified via participant self-reports. Prior adverse cardiac event was defined as any prior myocardial infarction, stroke, heart failure, or angina. Diabetes was defined as a participant who self-reported a diabetes diagnosis, were fasting with a glucose measurement of ≥7.00 mmol/L, or were not fasting with a glucose measurement of ≥11.10 mmol/L ^24^. Dyslipidemia was defined as a participant with a self-reported diagnosis of high cholesterol and/or total cholesterol of ≥6.21 mmol/L, HDL cholesterol of ≤1.19 mmol/L, LDL cholesterol of >4.1 mmol/L, or triglycerides >2.25 mmol/L. Hypertension was defined in participants with self-reported current use of anti-hypertensive medication and/or systolic blood pressure ≥140 mmHg, or diastolic blood pressure ≥90 mmHg ^25^. We also defined a binary variable indicating the presence of any of the following cardiometabolic conditions: diabetes, dyslipidemia, hypertension, or obesity.

### Statistical Analysis

We compared the distribution of sociodemographic variables, biomarkers, and comorbidities across the three PAD classifications: no PAD, clinical PAD, and borderline PAD. We then estimated the crude prevalence of clinical and borderline PAD across the total population and stratified across levels of each primary risk factor. To identify factors associated with clinical and borderline PAD in this setting, we calculated unadjusted and age-sex-adjusted prevalence ratios for each identified PAD risk factor. Prevalence ratios were estimated with log-binomial models and reported with 95% confidence intervals. We constructed separate models for clinical and borderline PAD. Clinical PAD cases were excluded from the borderline PAD analysis. All statistical analyses were conducted using R version 4.1.1 ^26^.

## Results

### Study Population

Our analytic cohort consisted of 1,883 HAALSI participants aged 40 to 69 years with complete ABI data. Over half (58.8%) of participants in this sample were female and approximately one-third (32.4%) had no formal education. Eighty-four percent of participants met the diagnostic criteria for at least one cardiometabolic disease and five percent reported a previous adverse cardiac event (see *Table 1*). Approximately 60% of the sample met the criteria for hypertension, 10% for diabetes, and 33% for obesity (see *Table S1*). Prevalence of hypertension and obesity was significantly greater in females compared to males.

**Table 1.** Sociodemographic characteristics and risk factors across PAD diagnostic categories.

|  | **No PAD**<br>(n=916) | **Clinical PAD**<br>(n=125) | **Borderline PAD**<br>(n=842) |
| --- | --- | --- | --- |
| Mean age, years | 53.97 (8.3) | 56.62 (7.8) | 53.05 (8.0) |
| Male, % | 45.6 (1.6) | 44.0 (4.4) | 36.0 (1.6) |
| Current smoker, % | 8.3 (0.9) | 10.4 (2.7) | 12.3 (1.1) |
| Current drinker, % | 19.1 (1.3) | 24.8 (3.9) | 20.5 (1.4) |
| Any CVD, % | 5.7 (0.8) | 7.2 (2.3) | 4.2 (0.7) |
| Any cardiometabolic disease, % | 84.0 (1.2) | 85.1 (3.2) | 83.2 (1.3) |
| Mean waist-to-hip ratio | 0.90 (0.1) | 0.92 (0.1) | 0.90 (0.1) |
| Mean BMI | 27.78 (6.6) | 27.88 (6.5) | 27.91 (7.3) |
| Mean waist circumference, cm | 93.64 (15.6) | 95.00 (16.2) | 92.39 (15.4) |
| Mean HDL, mmol/L | 1.55 (0.5) | 1.59 (0.6) | 1.58 (0.5) |
| Mean LDL, mmol/L | 2.09 (1.0) | 2.15 (1.1) | 2.02 (1.0) |
| Mean total cholesterol, mmol/L | 4.17 (1.2) | 4.35 (1.3) | 4.16 (1.2) |
| Mean triglycerides, mmol/L | 1.75 (0.9) | 1.84 (1.1) | 1.73 (0.9) |
| Mean glucose, mmol/L | 6.56 (2.9) | 7.11 (4.2) | 6.38 (3.1) |
| Mean diastolic BP, mmHg | 83.62 (12.1) | 85.08 (14.1) | 82.98 (11.8) |
| Mean systolic BP, mmHg | 136.10 (21.8) | 142.45 (24.2) | 133.36 (20.6) |
| Mean hemoglobin, g/dL | 12.59 (1.9) | 12.51 (1.9) | 12.63 (2.0) |
| Mean CRP, mg/L | 2.96 (2.9) | 3.92 (3.6) | 2.98 (3.1) |

### PAD Status

Overall, 6.6% (95% CI: 5.6-7.7) of the sample met the diagnostic criteria for clinical PAD while 44.7% (95% CI: 42.4-47.0) met the diagnostic criteria for borderline PAD (see *Table 2*). Clinical PAD was most prevalent among participants who were aged 60 to 69 years (9.3%, 95% CI: 6.9-12.1), current alcohol drinkers (8.2%, 95% CI: 5.6-11.4), had a high waist-to-hip ratio (9.8%, 95% CI: 5.5-15.9), had a previous adverse cardiac event (9.4%, 95% CI: 4.4-17.0), and diabetic (9.7%, 95% CI: 5.7-15.0). Borderline PAD was most prevalent among participants who were female (48.7%, 95% CI: 45.7-51.7), aged 40 to 49 years (48.7%, 95% CI: 44.6-52.8), and current smokers (53.6%, 95% CI: 46.3-60.8).

**Table 2.**
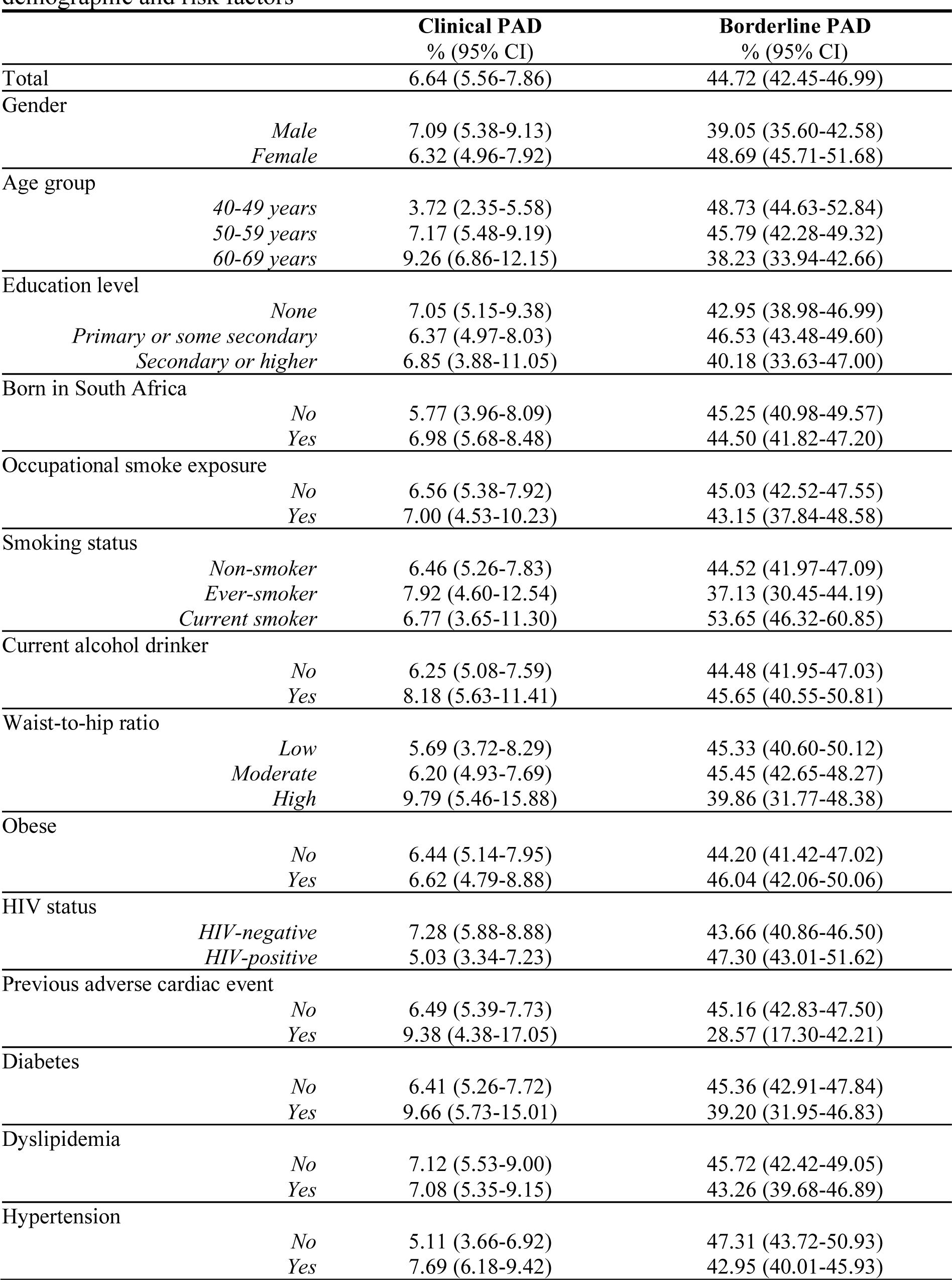
Estimated prevalence of clinical and borderline Peripheral Artery Disease, stratified by demographic and risk factors.

|  |  | Clinical PAD<br>% (95% CI) | Borderline PAD<br>% (95% CI) |
| --- | --- | --- | --- |
| Total |  | 6.64 (5.56-7.86) | 44.72 (42.45-46.99) |
| Gender | *Male* | 7.09 (5.38-9.13) | 39.05 (35.60-42.58) |
|  | *Female* | 6.32 (4.96-7.92) | 48.69 (45.71-51.68) |
| Age group | *40-49 years* | 3.72 (2.35-5.58) | 48.73 (44.63-52.84) |
|  | *50-59 years* | 7.17 (5.48-9.19) | 45.79 (42.28-49.32) |
|  | *60-69 years* | 9.26 (6.86-12.15) | 38.23 (33.94-42.66) |
| Education level | *None* | 7.05 (5.15-9.38) | 42.95 (38.98-46.99) |
|  | *Primary or some secondary* | 6.37 (4.97-8.03) | 46.53 (43.48-49.60) |
|  | *Secondary or higher* | 6.85 (3.88-11.05) | 40.18 (33.63-47.00) |
| Born in South Africa | *No* | 5.77 (3.96-8.09) | 45.25 (40.98-49.57) |
|  | *Yes* | 6.98 (5.68-8.48) | 44.50 (41.82-47.20) |
| Occupational smoke exposure | *No* | 6.56 (5.38-7.92) | 45.03 (42.52-47.55) |
|  | *Yes* | 7.00 (4.53-10.23) | 43.15 (37.84-48.58) |
| Smoking status | *Non-smoker* | 6.46 (5.26-7.83) | 44.52 (41.97-47.09) |
|  | *Ever-smoker* | 7.92 (4.60-12.54) | 37.13 (30.45-44.19) |
|  | *Current smoker* | 6.77 (3.65-11.30) | 53.65 (46.32-60.85) |
| Current alcohol drinker | *No* | 6.25 (5.08-7.59) | 44.48 (41.95-47.03) |
|  | *Yes* | 8.18 (5.63-11.41) | 45.65 (40.55-50.81) |
| Waist-to-hip ratio | *Low* | 5.69 (3.72-8.29) | 45.33 (40.60-50.12) |
|  | *Moderate* | 6.20 (4.93-7.69) | 45.45 (42.65-48.27) |
|  | *High* | 9.79 (5.46-15.88) | 39.86 (31.77-48.38) |
| Obese | *No* | 6.44 (5.14-7.95) | 44.20 (41.42-47.02) |
|  | *Yes* | 6.62 (4.79-8.88) | 46.04 (42.06-50.06) |
| HIV status | *HIV-negative* | 7.28 (5.88-8.88) | 43.66 (40.86-46.50) |
|  | *HIV-positive* | 5.03 (3.34-7.23) | 47.30 (43.01-51.62) |
| Previous adverse cardiac event | *No* | 6.49 (5.39-7.73) | 45.16 (42.83-47.50) |
|  | *Yes* | 9.38 (4.38-17.05) | 28.57 (17.30-42.21) |
| Diabetes | *No* | 6.41 (5.26-7.72) | 45.36 (42.91-47.84) |
|  | *Yes* | 9.66 (5.73-15.01) | 39.20 (31.95-46.83) |
| Dyslipidemia | *No* | 7.12 (5.53-9.00) | 45.72 (42.42-49.05) |
|  | *Yes* | 7.08 (5.35-9.15) | 43.26 (39.68-46.89) |
| Hypertension | *No* | 5.11 (3.66-6.92) | 47.31 (43.72-50.93) |
|  | *Yes* | 7.69 (6.18-9.42) | 42.95 (40.01-45.93) |

### Prevalence Ratios by Clinical PAD

Age and CRP were the only risk factors significantly associated with higher prevalence of clinical PAD (see *Figure 1*). After adjusting for gender, the 50-to-59-year age group was associated with nearly two times the prevalence of clinical PAD compared to the 40-to-49-year age group (PR: 1.9, 95% CI: 1.2-3.1) while the 60-to-69-year age group was associated with two-and-a-half times the prevalence of clinical PAD compared to the 40-to-49-year age group (PR: 2.5, 95% CI: 1.5-4.0). After adjusting for age and gender, the prevalence of clinical PAD increased by 8% for every 1 mg/L increase in CRP (PR: 1.08, 95% CI: 1.03-1.12).

**Figure 1.**
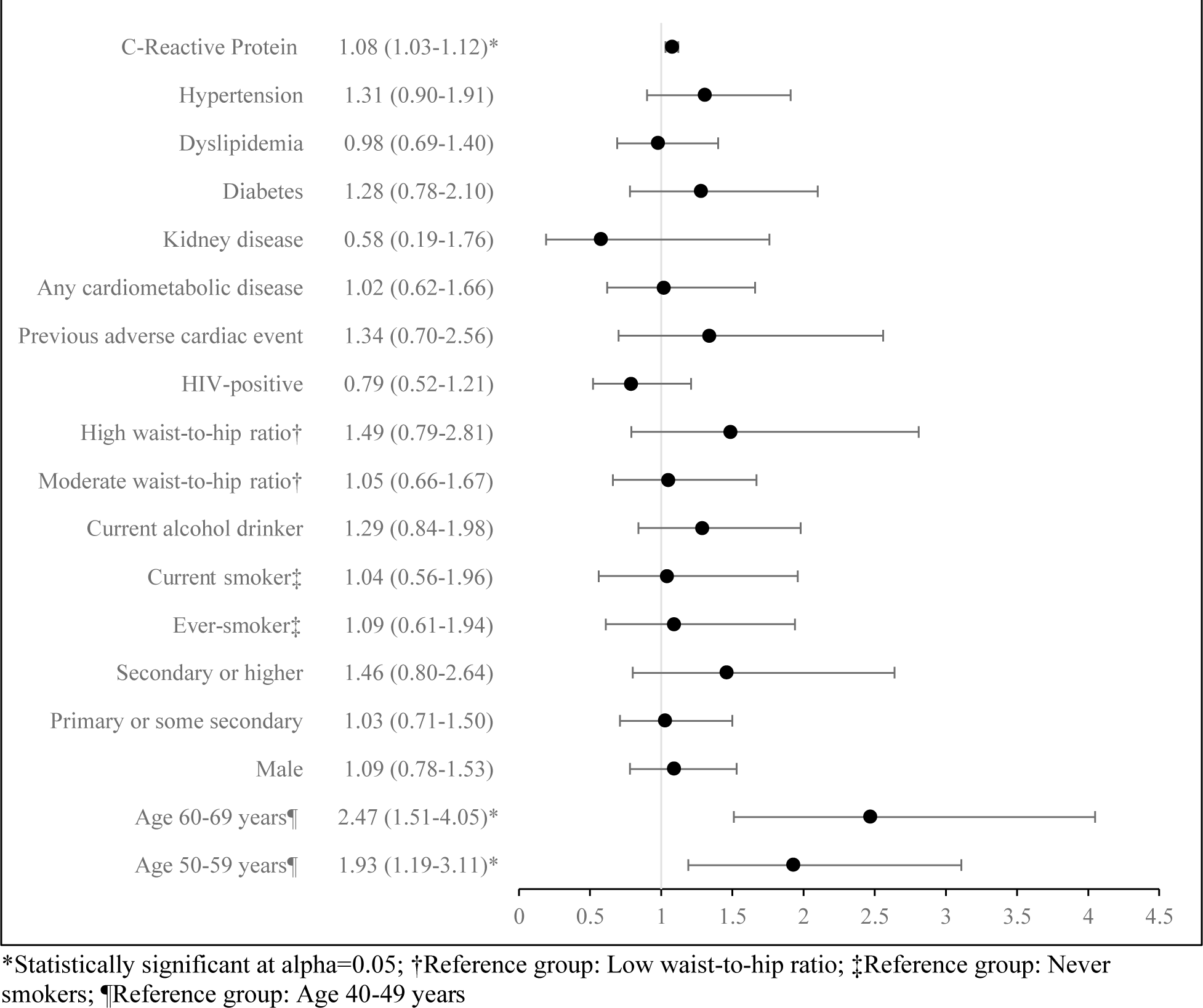
Age-sex adjusted prevalence ratios of clinical Peripheral Artery Disease

### Prevalence Ratios by Borderline PAD

Gender, age, current smoking status, and current drinking status were significantly associated with borderline PAD prevalence (see *Figure 2*). Males had 20% lower prevalence of borderline PAD compared to females, after adjusting for age (PR: 0.8, 95% CI: 0.7-0.9). The 60-to-69-year age group was also associated with an approximately 20% decrease in the prevalence of borderline PAD compared to the 40-to-49-year age group (PR: 0.8, 95% CI: 0.7-0.9). Being a current smoker (PR: 1.6, 95% CI: 1.3-1.9) and a current alcohol drinker (PR: 1.2, 95% CI: 1.0-1.3) were each independently associated with higher prevalence of borderline PAD after adjusting for age and gender.

**Figure 2.**
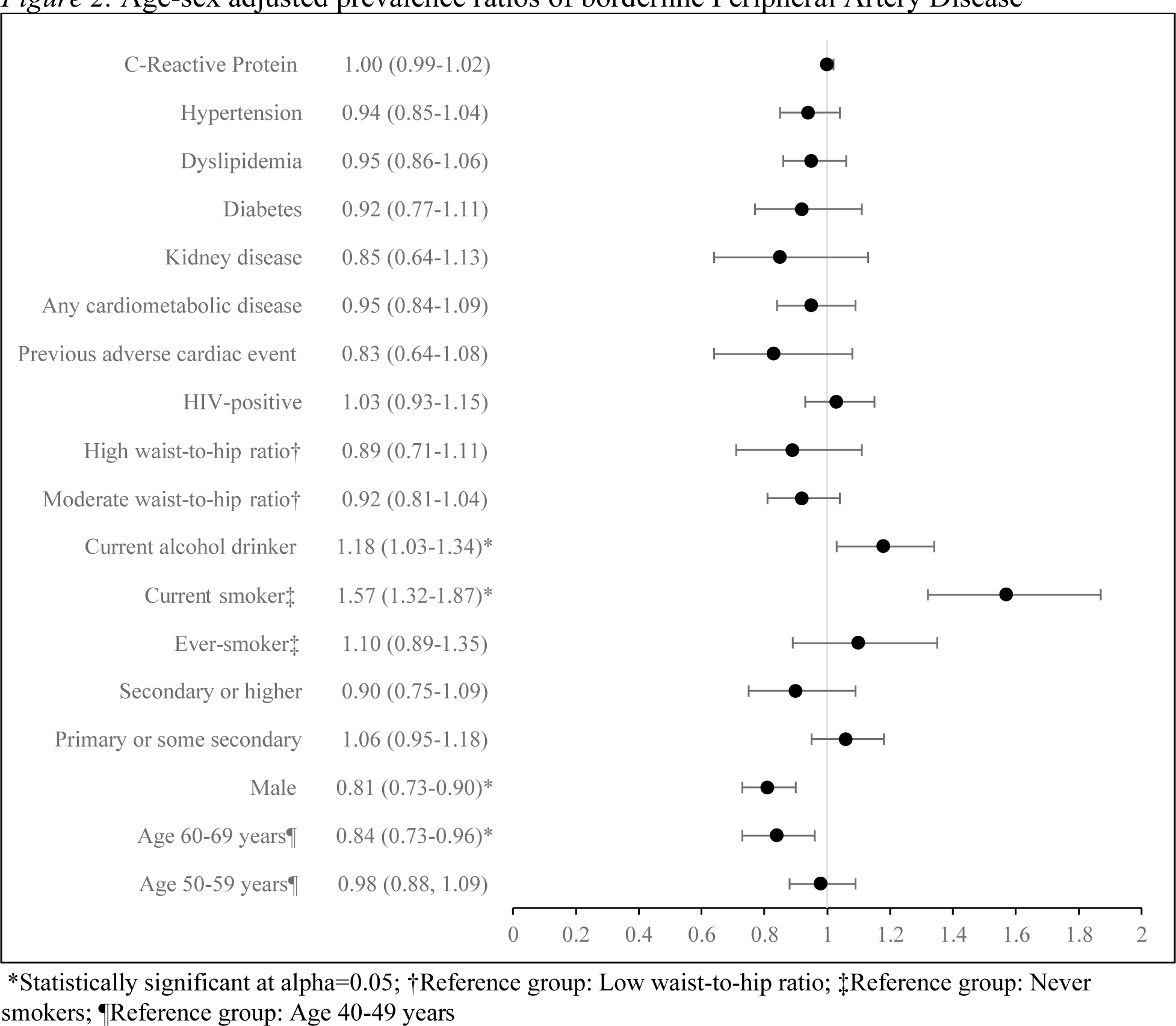
Age-sex adjusted prevalence ratios of borderline Peripheral Artery Disease

## Discussion

We conducted an analysis of the HAALSI cohort and provide the first estimate of PAD prevalence for a rural South African population. We estimated that the prevalence of clinical PAD in this representative sample of adults aged 40 to 69 years was 6.6%. Also, despite an average age of 54 years, nearly half of our sample met the diagnostic criteria for borderline PAD. Most traditional risk factors (e.g., smoking, diabetes, hypertension, and hypercholesterolemia) were similar across strata of PAD. Age and CRP were the only factors significantly associated with clinical PAD while age, gender, current smoking status, and current alcohol drinking status were significantly associated with borderline PAD.

Our estimate of clinical PAD prevalence was lower than previous studies from sub-Saharan Africa which range from 14% to 38% ^9,10,12,27^. However, these studies focused primarily on populations older than 60 years while our population was between 40 and 69 years old. In contrast, our prevalence measures were higher than prior estimates from LMIC settings performed in similar age groups. Studies from the Philippines, Malawi, and China estimated a prevalence of PAD ranging from 2% to 5% for comparable age groups ^28–31^. Similarly, a previous global analysis concluded that LMICs have higher PAD prevalence at younger ages compared to high-income countries ^7^. We estimated the prevalence of clinical PAD in HAALSI participants aged 40-49 as well as aged 50-59 to be over two times higher than estimates from the United States ^32^. This high prevalence implies the need for early intervention to slow and reduce PAD risk for South African adults.

We found that nearly half of HAALSI participants that were assessed for PAD had a borderline ABI measurement between 0.91 and 1.00. In contrast, borderline PAD prevalence has been measured at between 9% and 17% across a variety of settings ^30,33–36^. Borderline ABI is a sign of premature atherosclerotic disease and is associated with higher risk of coronary heart disease, adverse vascular events in the leg, and progression to clinical PAD compared to normal ABI ^36,37^. Therefore, without proper identification and management, a significant percent of this rural South African population may be at high risk for future disease progression and adverse cardiovascular events. This risk is even more pronounced given that over 80% of the population also suffers from at least one cardiometabolic disease. Future research should assess the benefits of early PAD detection for high-risk groups in LMIC settings as a strategy to reduce downstream adverse cardiovascular events and premature death.

PAD risk factors may differ in sub-Saharan Africa compared to high-income countries. For example, PAD has been found to be significantly higher among females and associated with both underweight and obesity ^9–11,38^. This is different from high-income settings, like the United States, where there are little gender differences in PAD prevalence ^7^. The gender differences are particularly interesting considering that traditional risk factors, such as smoking, tend to be lower among females in LMIC settings ^7^. Though, high rates of obesity and hypertension were found in the female HAALSI population which may contribute to the development of PAD. We did not find that traditional risk factors (diabetes, hypertension, dyslipidemia, smoking, alcohol drinking, and obesity) were associated with a higher likelihood of having clinical PAD. In fact, only high CRP and age were associated with PAD in our study. The association between CRP and PAD is supported by previous studies from high-income countries ^7,39^. We also found that being a current smoker or current alcohol drinker was associated with borderline PAD status, findings in line with prior literature ^5,7^. Together, this suggests that unknown underlying risk factors are likely influencing the high rates of PAD observed in the HAALSI population.

Alternative potential risk factors for PAD in this population include endemic infectious diseases, chronic stress, and air pollution. Preliminary ecological evidence shows an association between PAD prevalence and country-level burdens of endemic infectious diseases like HIV, tuberculosis, and malaria ^40^. Historically, rural South Africa has had a high infectious disease burden, including HIV. Approximately 23% of the HAALSI population is HIV-positive with South Africa having one of the highest HIV burdens globally ^16^. Additionally, chronic stress may be a critical factor in the formation of atherosclerosis via the impacts of chronic inflammation ^41^. The historical context of apartheid and the low-income, high unemployment environment of the Agincourt sub-district has likely led to high levels of lifetime stress for HAALSI participants.

This high stress environment may be contributing to the early development of PAD indicated by the prevalence of borderline ABIs. Finally, associations have been observed between long-term exposure to air pollutants, including carbon monoxide and particulate matter, and PAD ^42^. While outdoor air pollution is not substantial in Agincourt, HAALSI participants may have been exposed to high levels of indoor air pollutants from indoor cooking with fire as observed in similar settings elsewhere ^28^. These risk factors warrant future exploration in the HAALSI cohort.

As the only factor other than age associated with clinical PAD in this study, CRP demonstrates the possible importance of life stressors and inflammation in the development of PAD and other atherosclerotic conditions. Chronic stress has been identified as a risk factor for atherosclerosis, particularly through its link to chronic inflammation ^43^. CRP is one measure of inflammation associated with cardiovascular disease that is likely influenced by both acute infections and stress exposure over the life course ^44–46^. This setting of rural South Africa is a population with both high exposure to chronic stressors and high infectious disease burdens. A high proportion of the HAALSI population has experienced one or more traumatic events throughout the life course including severe financial hardship, life-threatening illnesses, death of a close family member/friend from violence, and physical abuse by parents ^47^. The underlying history of apartheid may also contribute to this, creating a unique environment characterized by high stress across the life course.

The primary limitation of this study is its cross-sectional nature. Because of this design, we cannot confirm the temporal link between the examined risk factors and the development of PAD. However, future work can exploit the additional waves of data collection from the HAALSI cohort to create a more nuanced picture of PAD development and progression for this population. Additionally, only a sub-sample of HAALSI participants were included in this analysis and no one above the age of 69 was included. Therefore, this analysis is not representative of the total population affected by PAD and primarily describes cases with early onset. However, a comparison of key sociodemographic characteristics in the full HAALSI sample of individuals aged <70 years to the subset used in this analysis showed few differences. Due to the smaller sample size compared to the total HAALSI population, our study may not be powered appropriately to detect significant differences across all traditional risk factors. Despite these limitations, our analysis is strengthened by the use of the ABI as an objective diagnostic measure for PAD.

## Conclusion

Our study was the first to quantify the PAD burden and profile potential risk factors in a rural South African population. Most notably, we found that nearly half of HAALSI participants met the diagnostic criteria for borderline PAD. As the population of this study ranged from 40 to 69 years, this represents a large proportion with borderline disease at a younger age than has been previously documented in the literature. We also found that age and CRP were the only factors significantly associated with clinical PAD. No other traditional risk factors were found to significantly predict clinical PAD in this population. Future research should focus on identifying the unknown, underlying risk factors that influence the high rates of PAD observed in this population. South African policymakers and clinicians should consider expanded screening for early PAD detection in rural areas.

## Non-standard Abbreviations and Acronyms

Health and Aging in Africa: A Longitudinal Study of an INDEPTH Community (HAALSI) Health and Demographic Surveillance System (HDSS)

## Data Availability

The data underlying this article are available in the Harvard Dataverse at https://doi.org/10.7910/DVN/TW84UI & https://doi.org/10.7910/DVN/F5YHML.

https://doi.org/10.7910/DVN/TW84UI

## Acknowledgements

The authors have no acknowledgements.

## Sources of Funding

The authors did not receive support from any organization for the submitted work.

## Disclosures

The authors have no disclosures to declare that are relevant to the content of this article.

## Supplemental Material

Table S1

---

**Table S1.** Prevalence of comorbid chronic conditions, stratified by gender.

|  | **Total Sample**<br>% (95% CI) | **Male**<br>% (95% CI) | **Female**<br>% (95% CI) |
| --- | --- | --- | --- |
| Obesity | 33.26 (31.12-35.45) | 15.75 (13.23-18.53) | 45.40 (42.43-48.4) |
| HIV status | 30.51 (28.37-32.72) | 30.18 (26.84-33.68) | 30.74 (27.95-33.64) |
| Diabetes | 9.87 (8.53-11.35) | 8.48 (6.56-10.74) | 5.89 (4.55-7.49) |
| Dyslipidemia | 45.45 (43.02-47.89) | 45.91 (42.06-49.80) | 45.14 (42.01-48.31) |
| Hypertension | 59.18 (56.91-61.42) | 53.32 (49.72-56.89) | 63.27 (60.35-66.13) |

## Notes

### Competing Interest Statement

The authors have declared no competing interest.

### Author Declarations

The HAALSI study has ethical approvals from the Mpumalanga Provincial Research and Ethics Committee, the University of the Witwatersrand Human Research Ethics Committee (ref. M141159), and the Harvard T.H. Chan School of Public Health Office of Human Research Administration (ref. C13-1608-02). Ethical approvals were also sought from the Indiana University Institutional Review Board for this analysis where it was deemed not human subjects research because analysis was conducted on deidentified datasets (ref. 18808). Informed consent was obtained from all individual participants included in the study.

